# Anticipating conversion to hysterectomy in women counseled for myomectomy: temporal validation of the *EFFAH* score at a Ghanaian district hospital

**DOI:** 10.64898/2026.10.26.26366536

**Authors:** Kofi Effah, Nana Owusu Mensah Essel, Joseph Emmanuel Amuah, Abraham Kwadzo Ahiakpa, Christopher Nyamekye, Stephen Danyo, Eli Yao Lawluvi, Hannah Venunye Dzefe, Augustina Ayinpoka Asolmia, Frank Aformanya, Joseph Ofori Pabby, Eugene Owuraku Koranteng Atiemo, Bernard Hayford Atuguba

**Affiliations:** Department of Obstetrics and Gynaecology, Catholic Hospital, Battor, P. O. Box 2, Battor Via Sogakope, Volta Region, Ghana; Cervical Cancer Prevention and Training Centre, Catholic Hospital, Battor, P. O. Box 2, Battor Via Sogakope, Volta Region, Ghana; School of Public Health, College of Health Sciences, University of Alberta, Edmonton, Alberta, Canada; Department of Emergency Medicine, Faculty of Medicine & Dentistry, College of Health Sciences, University of Alberta, Edmonton, Alberta, Canada; School of Epidemiology and Public Health, University of Ottawa, Ottawa, Canada; Department of Internal Medicine, Ho Teaching Hospital, P. O. Box MA 374, Ho, Volta Region, Ghana; Department of Obstetrics and Gynaecology, Korle Bu Teaching Hospital, Accra, Ghana; Department of Internal Medicine, Catholic Hospital, Battor, P. O. Box 2, Battor Via Sogakope, Volta Region, Ghana; Centre for Plant Medicine Research, P. O. Box 73, Mampong-Akuapem, Ghana; Sacred Heart Catholic Hospital, Abor-Anyako Main Road, P. O. Box AB14, Weme Abor, Volta Region, Ghana; Department of Surgery, Komfo Anokye Teaching Hospital, Kumasi, Ghana

**Keywords:** uterine fibroids, myomectomy, hysterectomy, fertility preservation, clinical prediction rule, validation study

## Abstract

**Background:** In women with uterine fibroids counseled for myomectomy, consent typically involves discussing the preoperative probability that the surgery will end in hysterectomy. To our knowledge, no published tool estimates this probability, including in sub-Saharan Africa. This study aimed to temporally validate the Estimating Fibroid Factors Associated with Hysterectomy (*EFFAH*) score (an eight-item bedside score) in a later cohort at a district hospital in Ghana, to define risk groups for counseling and triage.

**Methods:** In this retrospective cohort study with temporal external validation, the development cohort comprised 392 records of women counseled for myomectomy with possible hysterectomy who underwent surgery at Catholic Hospital, Battor, in 2013 to 2014. A pilot analysis was conducted in July 2025 to arrive at the odds ratio used to calculate a validation sample size of 416. The validation cohort comprised 800 women counseled for myomectomy with possible hysterectomy in 2017 to 2019. The score assigns points to eight items from the history, abdominal examination, hemoglobin and renal ultrasound. We assessed discrimination, calibration and net benefit of the development equation with 95% confidence intervals (CIs), updated it by closed testing and compared it with AutoScore, a random forest-based points score.

**Results:** Hysterectomy was performed in 31.6% of women in the development cohort and 21.1% in the validation cohort. The *C*-statistic was 0.76 (95% CI: 0.70, 0.80) at development and 0.73 (95% CI: 0.69, 0.77) at validation. The development equation overestimated risk, with an observed-to-expected ratio of 0.69 (95% CI: 0.60, 0.78) and a calibration slope of 0.83 (95% CI: 0.63, 1.03), and an intercept-only update was selected. The observed proportion with hysterectomy was 6.2% for scores of 0 to 2, 23.5% for 3 to 4 and 42.2% for ≥5; a score below 3 yielded a negative predictive value of 93.8% (95% CI: 90.1, 96.2). AutoScore yielded a C-statistic of 0.74 (95% CI: 0.69, 0.78) and did not outperform the score (difference 0.01; 95% CI: −0.02, 0.04).

**Conclusions:** The *EFFAH* score ranked women by their risk of hysterectomy in a later cohort at the same hospital, and an intercept update aligned its average predicted risk with the observed proportion in that cohort. Clinically, about 6 in 100 women scoring 0 to 2 and about 4 in 10 women scoring ≥5 underwent hysterectomy; for the latter, explicit consent for hysterectomy, a senior surgeon and readily available cross-matched blood are proposed. Because it uses only routine preoperative findings, the score could support consent and resource planning in resource-limited and resource-rich settings alike, after local recalibration and prospective multicenter evaluation.

## Introduction

Leiomyomas are the most common benign tumors of the female reproductive system; their global incidence increased from 6.0 million in 1990 to 10.1 million in 2021, with the burden shifting toward countries with a low sociodemographic index^1^. Among Black women, cumulative prevalence by 50 years exceeds 80%^2^, two to three times the risk in white women^3^. In sub-Saharan Africa, women often present late with a large uterus; 88.7% of 300 women in Lagos had a uterus of ≥20 weeks at presentation^4^, and transfusion at abdominal myomectomy ranges from 22.9% to 58.2% in Nigerian series^5–7^. Hysterectomy is often culturally unacceptable because of the value placed on childbearing and the uterus^8–10^, and myomectomy is the fertility-sparing option^11^. For a woman who wishes to preserve her uterus, counseling and consent address myomectomy with possible hysterectomy. The preoperative probability of hysterectomy is therefore central to informed consent. Women who desired fertility but underwent hysterectomy have reported worse quality of life afterwards^12^, and women with fibroids report wanting complete information before surgery^13^.

To the best of our knowledge, no published tool estimates this probability before surgery. In Nigeria, surgeons have recommended consent for hysterectomy before myomectomy^14^ and have obtained consent for open myomectomy with possible hysterectomy when fibroids are very large^15^. Guidance from the Society of Obstetricians and Gynaecologists of Canada similarly states that women should be counseled about the risk of requiring a hysterectomy at a planned myomectomy but provides no estimate^16^. The practice bulletin of the American College of Obstetricians and Gynecologists on fibroids similarly recommends shared decision-making and myomectomy for women who want uterine preservation without addressing consent for hysterectomy^17^. Reported frequencies further seem to vary with setting and case mix; 3.0% of 13,213 myomectomies in a United States registry underwent a hysterectomy under the same anesthetic^18^, 1.0% of 35,151 abdominal myomectomies in California were converted to hysterectomy^19^, and 6% of repeat abdominal myomectomies for large uteri in the U.K. ended in hysterectomy for bleeding^20^. The registry analysis reported associations with age and operative time instead of a prediction model, and its authors could not determine whether the hysterectomies were planned^18,21^. In Nigeria, hysterectomy followed one of 276 abdominal myomectomies in Ibadan^22^, two of 224, both at re-operation for bleeding^23^, and two planned myomectomies in a Kano series of 105 women^14^. Existing prediction tools in fibroid surgery address transfusion at myomectomy^24,25^, conversion of laparoscopic to open myomectomy^26^ or conversion and complications at hysterectomy^27–29^. All were developed in high-resource settings, several include surgical route or operative time as predictors, and none was developed for a district hospital or limited-resource setting where myomectomy may be performed by open surgery and the uterus is often at or above the umbilicus at presentation.

The Estimating Fibroid Factors Associated with Hysterectomy (hereafter *EFFAH* score) was developed at Catholic Hospital, Battor, a district hospital in the Volta Region of Ghana. It sums points for eight items recorded at the preoperative assessment: age, parity, uterine size on abdominal examination, previous myomectomy, previous laparotomy, hydronephrosis on the renal scan performed with the routine pelvic ultrasound, hemoglobin concentration and past transfusion. Uterine size, low hemoglobin concentration, previous myomectomy and previous abdominal surgery are established markers of bleeding, transfusion or hysterectomy in fibroid surgery^7,22,30–33^. Hydronephrosis becomes more frequent as the uterus enlarges^34^, and older age was found to be the strongest preoperative factor associated with hysterectomy at myomectomy in a United States registry^18^. Evaluation of a prediction score in patients outside the development cohort requires assessment of calibration and net benefit as well as discrimination^35–37^. Whether a machine learning model would outperform a simple points score is also an empirical question; a systematic review found no such advantage in most clinical settings^38^.

This study aimed to validate the *EFFAH* score for predicting hysterectomy among women counseled for myomectomy with possible hysterectomy in a later cohort at the same hospital and to update its equation for that period. The secondary aim was to compare the *a priori* score (whose points were allocated based on expert opinion) with AutoScore, a random forest-based points score built from the same items.

## Methods

### Study design and reporting

This was a retrospective cohort study with temporal external validation at a single center. The score was first analyzed as a pilot in the development cohort, which informed the sample size, and was then evaluated without changing its items or points in a later validation cohort whose records were extracted from patient charts and theater registers. We report the study in accordance with the TRIPOD+AI statement, the update of the Transparent Reporting of a multivariable prediction model for Individual Prognosis Or Diagnosis (TRIPOD) guideline for regression and machine learning models^35^.

### Setting and participants

Both cohorts were drawn from the Department of Obstetrics and Gynaecology, Catholic Hospital, Battor, North Tongu District, Volta Region, Ghana, a district hospital that provides gynecologic surgery and serves as a major referral and training center^39–42^. In both cohorts, women were eligible if they had been counseled preoperatively for myomectomy with possible hysterectomy for uterine fibroids. The development cohort comprised all 392 eligible surgical records from 2013 and 2014. For the validation cohort, the study team screened 1,132 records of women who underwent surgery between 2017 and 2019. Records were excluded if the woman had been counseled for hysterectomy only, if the operation was also performed for another indication, such as a pelvic abscess, or if the record duplicated another; records without an eligibility status were also excluded (**Figure 1**). The study was approved by the Ethics Committee of Catholic Hospital, Battor (CHB-ERC-002/01/24), which also waived the need for individual consent on the basis that this was a retrospective analysis of anonymized records.

**Figure 1.**
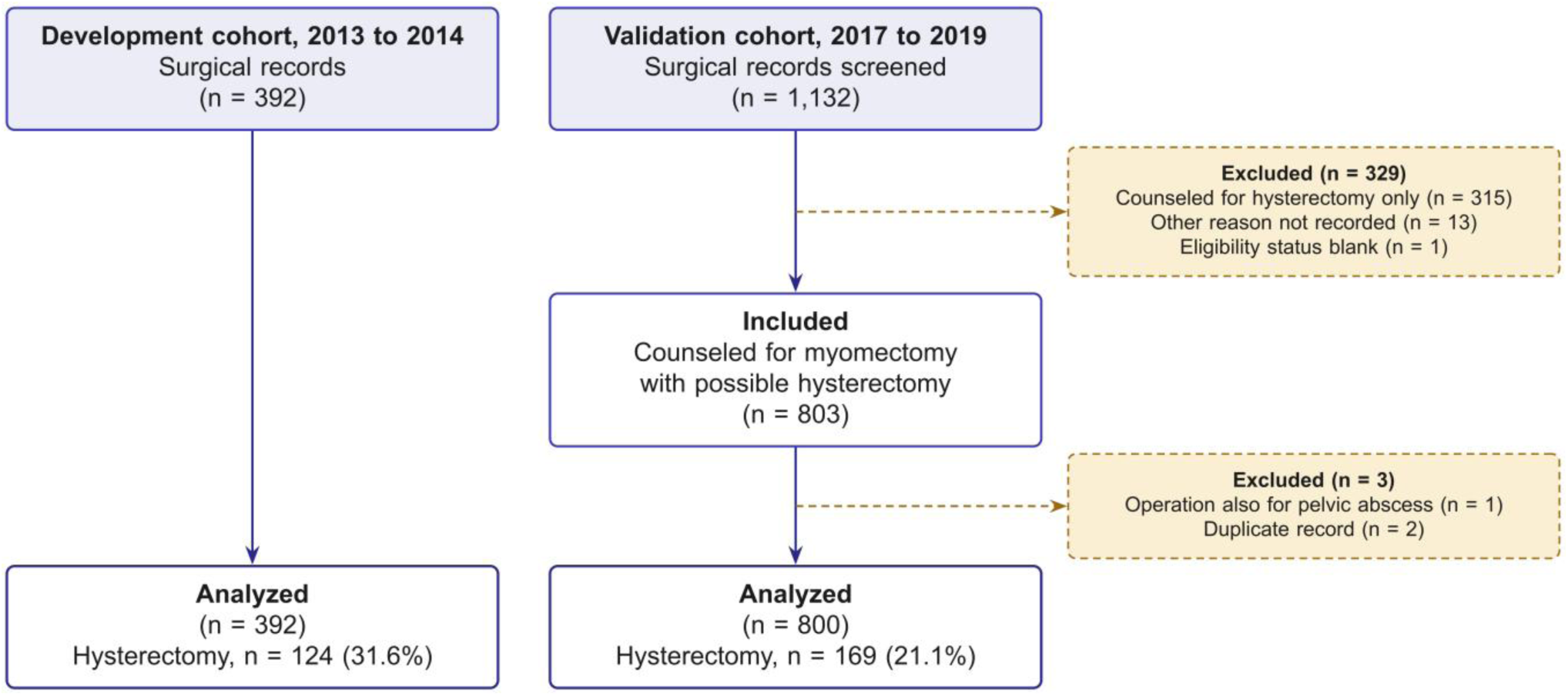
Selection of records in the development (2013 to 2014) and validation (2017 to 2019) cohorts.

### Outcomes

The primary outcome was hysterectomy (total or subtotal abdominal hysterectomy) at the index surgery, as coded by the study team from the operation record. The outcome therefore reflects operative findings (the surgeon and the woman’s consent) as well as lesion severity (indirectly). Secondary outcomes were estimated blood loss, intraoperative blood transfusion and relaparotomy, taken from the complications section of the theater register.

### The *EFFAH* score

The eight items and their points are shown in **Figure 2A**; the points were fixed a priori by expert opinion and were not re-estimated in either cohort. Two items were scored differently from the original scoring form because of how they were recorded in the data. First, the form assigns 2 points for one previous myomectomy and 3 points for two previous myomectomies, but the records stated only whether a previous myomectomy had taken place; any previous myomectomy was therefore assigned 2 points, and the maximum total was 15 instead of 16. Second, the form specifies hemoglobin at the first encounter with the patient, whereas both cohorts were scored on the last preoperative hemoglobin, the value used in the pilot analysis; hemoglobin at first encounter was examined in a sensitivity analysis.

**Figure 2.**
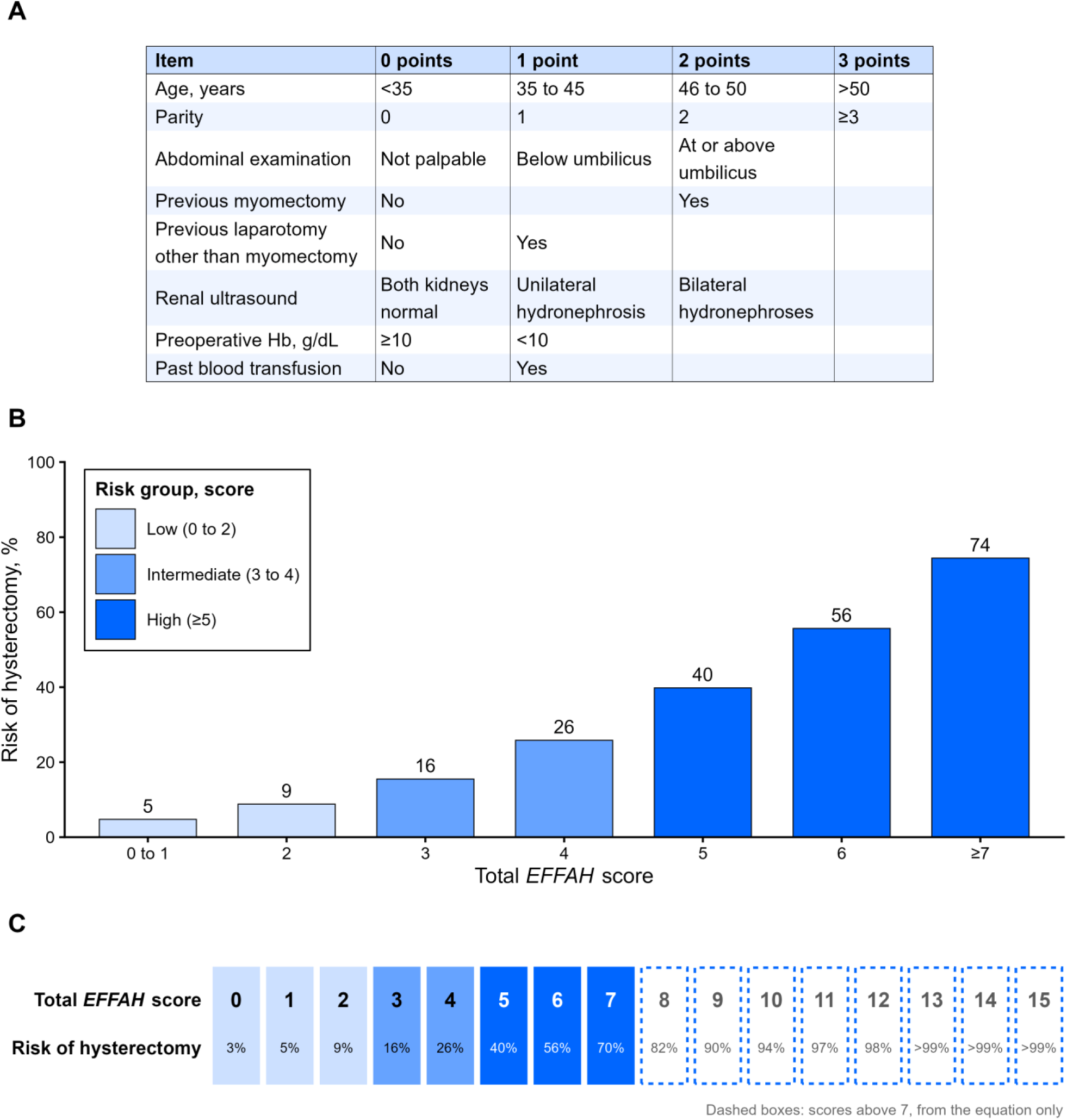
*EFFAH* score card. (A) Items and points. (B) Risk of hysterectomy by total score in the validation cohort, from the equation updated to 2017 to 2019 (apparent). (C) Predicted risk at each total score from the same equation. Dashed boxes mark scores above 7, for which risks come from the equation only because these scores were pooled with 7 in the validation analysis. Abbreviations: *EFFAH*, Estimating Fibroid Factors Associated with Hysterectomy; Hb, hemoglobin.

### Risk groups and proposed clinical actions

Three risk groups were defined from the development cohort at the score values where the observed proportion with hysterectomy increased most steeply: low (0 to 2 points), intermediate (3 to 4) and high (≥5). We linked each group to a proposed clinical action (**Table 1**), i.e., counseling with the frequency of hysterectomy for the low-risk group; counseling and consent for possible hysterectomy for the intermediate-risk group; and a senior surgeon, readily available cross-matched blood and consideration of referral for the high-risk group. The groups were chosen after the observed proportions in both cohorts had been inspected, and the actions are proposed uses of the score that were not evaluated in this study.

**Table 1.** Hysterectomy by *EFFAH* risk group in the development (n = 392) and validation (n = 800) cohorts.

| Risk group, score | Proposed action | Development, n | Development observed, % (95% CI) | Validation, n | Validation observed, % (95% CI) | Adjusted risk for counseling, % | Development complete cases, k/n; % (95% CI) | Validation complete cases, k/n; % (95% CI) |
| --- | --- | --- | --- | --- | --- | --- | --- | --- |
| Low (0 to 2) | Counsel that about 8 in 100 undergo hysterectomy | 133 | 9.1 (5.0, 15.9) | 266 | 6.2 (3.8, 9.9) | 7.7 | 6/61; 9.8 (4.6, 19.8) | 13/206; 6.3 (3.7, 10.5) |
| Intermediate (3 to 4) | Counsel and consent for possible hysterectomy | 175 | 33.4 (26.4, 41.2) | 389 | 23.5 (19.5, 28.1) | 19.8 | 27/100; 27.0 (19.3, 36.4) | 67/294; 22.8 (18.4, 27.9) |
| High ($\geq 5$) | Senior surgeon, readily available cross-matched blood, consider referral | 83 | 63.8 (52.3, 73.9) | 145 | 42.2 (34.2, 50.6) | 49.3 | 21/37; 56.8 (40.9, 71.3) | 41/102; 40.2 (31.2, 49.9) |
| Abbreviations: CI, confidence interval; *EFFAH*, Estimating Fibroid Factors Associated with Hysterectomy.<br>n is the mean group size across 20 imputations. Adjusted risk is from the updated equation. Complete cases: women with all eight items recorded. Complete-case 95% CIs use the Wilson score method. |  |  |  |  |  |  |  |  |

### Sample size

The sample size for the validation cohort was calculated in July 2025 from the pilot results with G*Power 3.1.9.7 (Heinrich Heine University Düsseldorf, Düsseldorf, Germany)^43^. Using the two-tailed Wald z test for a logistic regression coefficient with an odds ratio (OR) of 1.89 per score point, a probability of hysterectomy of 0.2 under the null hypothesis, a two-sided alpha of 0.05, a power of 0.8 and a binomial predictor distribution, the required sample size was determined to be n = 416. Because this calculation treats the score effect as a single contrast, we also report the precision achieved for the C-statistic, the observed-to-expected (O/E) ratio and the calibration slope using the formulas put forward by Riley et al. for external validation^44,45^. No sample size was calculated for the development cohort, which comprised all eligible records from 2013 and 2014.

### Missing data

Missing score items were handled via multiple imputation by chained equations^46,47^. For the development cohort, we used the 20 imputed datasets created for the pilot (*mice* default methods, 10 iterations). For the validation cohort, we imputed missing items with the same approach (20 datasets, 10 iterations; predictive mean matching for parity and hemoglobin, logistic regression for binary items and polytomous regression for abdominal examination and renal ultrasound), with the outcome and the number of living children in the imputation model. Items were imputed as raw values, and the score was calculated after imputation. Estimates were combined with Rubin’s rules; C-statistics and proportions were pooled on the logit scale^46^. The imputation diagnostics are reported in **Table S1 and Figures S1 and S2**.

### Statistical methods

#### Development cohort

In each imputed dataset, we fitted a logistic regression of hysterectomy on the total score and pooled the coefficients with Rubin’s rules. The pooled intercept and slope form the development equation, which converts a total score into a predicted probability. A logistic regression with the eight items as separate predictors was fitted in the same way to show the contribution of each item (**Table S2**). Linearity of the log odds in the total score was tested by comparing a restricted cubic spline with three knots with the linear term, using a likelihood ratio test pooled across imputations (**Table S3**). Collinearity among the eight items was assessed with generalized variance inflation factors adjusted for the degrees of freedom of each item. Influential records were identified with Cook’s distance and with the standardized change in the score coefficient when each record was omitted (dfbetas).

#### Validation and updating

We applied the development equation unchanged to the validation cohort. Discrimination was assessed with the C-statistic. Calibration was assessed with the calibration intercept (calibration in the large), the calibration slope, the O/E ratio, the Brier score and a flexible calibration curve^36,48^. Because the proportion with hysterectomy differed between the two periods, the equation was updated to the validation cohort using the closed testing procedure described by Vergouwe et al.^49^. This procedure compares three nested models: the unchanged development equation; recalibration in the large, which re-estimates the intercept while the coefficient of the linear predictor is fixed at 1; and logistic recalibration, which re-estimates both the intercept and the coefficient of the linear predictor. In each imputed dataset, we fitted the recalibration model *logit*(*p*) = *a* + *b* × *LP*, where *LP* is the linear predictor of the development equation, and pooled a and b with Rubin’s rules. The joint hypothesis *a* = 0 and *b* = 1 was tested with a pooled Wald test (D1 statistic); if it was rejected, the hypothesis b = 1 was tested, and recalibration in the large was retained unless this second test was also rejected, with alpha 0.05 for both tests (**Table S3**). Performance estimates of the updated equation in the validation cohort are considered apparent; optimism was estimated with 1,200 bootstrap samples that repeated the entire testing and updating procedure. The updated equation gives the adjusted risks in **Table 1** and **Figure 2**. Two sensitivity analyses scored hemoglobin at first encounter, as specified on the form, and restricted the analysis to women with all eight items recorded (complete cases) (**Table S4**).

#### Risk groups and clinical utility

For each risk group, we report the proportion with hysterectomy in both cohorts and, for the cut-offs at 3 and 5 points, the sensitivity, specificity and positive and negative predictive values (**Table S5**). Decision curve analysis estimated the net benefit of using the development equation to identify women at high risk of hysterectomy, across threshold probabilities of 10% to 40%, compared with treating all or no women as high risk^37,50,51^. Estimated blood loss (median and interquartile range [IQR]), intraoperative transfusion and relaparotomy were summarized by risk group in each cohort (**Table S6**).

#### AutoScore comparison

As a secondary aim, the score was compared with a points score built with AutoScore^52^, a framework that generates integer risk scores from routinely collected data. AutoScore was developed in the first imputed development dataset, which was split by folder number into training (60%), validation (20%) and test (20%) sets. The eight items were ranked by random forest variable importance (100 trees), and all eight were retained. Age, parity and hemoglobin were categorized at their 5th, 20th, 80th and 95th percentiles, after which the parity cut-off was set at 1 birth and the hemoglobin cut-off at 9 g/dL on clinical grounds. A logistic regression of hysterectomy on the categorized items in the training set gave coefficients that were converted to integer points, scaled so that the maximum total was 50. The total points were converted to a probability by logistic regression in the development dataset. The points table and this equation were applied unchanged to each validation imputation, and the C-statistics of AutoScore and the *EFFAH* score were compared with the paired DeLong test, pooled across imputations (**Table S7**).

All statistical analyses were conducted using R version 4.6.1 (R Foundation for Statistical Computing, Vienna, Austria) with the packages *targets* 1.12.0, *mice* 3.19.0, *pROC* 1.19.1, *car* 3.1.5, *randomForest* 4.7.1.2 and *AutoScore* 1.1.0. Hypothesis testing was performed at a two-tailed alpha level of 5%.

## Results

### Participants and development cohort

The development cohort comprised 392 records, of which 124 (31.6%) ended in hysterectomy, and the validation cohort comprised 800 records, of which 169 (21.1%) ended in hysterectomy (**Figure 1**). Case mix was similar in the two periods (**Table 2**). The median age was 37 years in both cohorts (IQR, 33–40 and 33–41), most women were nulliparous, and the uterus was palpable at or above the umbilicus in 58.8% of development and 60.2% of validation records. Previous laparotomy (6.4% and 9.1%), past transfusion (13.7% and 18.0%) and preoperative hemoglobin below 10 g/dL (10.6% and 13.3%) were slightly more frequent in the validation cohort. Renal ultrasound was missing for 177 (45.2%) development and 142 (17.8%) validation records, and each other item was missing for less than 7% of records (**Table S1**).

**Table 2.** Characteristics of women undergoing surgery for uterine fibroids, development and validation cohorts.

| **Characteristic** | **Development, 2013 to 2014 (n = 392)** | **Development missing, n** | **Validation, 2017 to 2019 (n = 800)** | **Validation missing, n** |
| --- | --- | --- | --- | --- |
| Hysterectomy, n (%) | 124 (31.6) | 0 | 169 (21.1) | 0 |
| Age, years; median (IQR) | 37 (33 to 40) | 0 | 37 (33 to 41) | 0 |
| Parity; median (IQR) | 0 (0 to 1) | 10 | 0 (0 to 1) | 35 |
| Abdominal examination, n (%) |  | 18 |  | 19 |
| Uterus not palpable | 27 (7.2) |  | 24 (3.1) |  |
| Palpable below umbilicus | 127 (34.0) |  | 287 (36.7) |  |
| Palpable at or above umbilicus | 220 (58.8) |  | 470 (60.2) |  |
| Previous myomectomy, n (%) | 16 (4.3) | 22 | 37 (4.7) | 15 |
| Previous laparotomy other than myomectomy, n (%) | 24 (6.4) | 18 | 72 (9.1) | 11 |
| Renal ultrasound, n (%) |  | 177 |  | 142 |
| Both kidneys normal | 200 (93.0) |  | 622 (94.5) |  |
| Unilateral hydronephrosis | 9 (4.2) |  | 18 (2.7) |  |
| Bilateral hydronephroses | 6 (2.8) |  | 18 (2.7) |  |
| Preoperative Hb, g/dL; median (IQR) | 11.6 (10.7 to 12.6) | 5 | 11.4 (10.5 to 12.4) | 47 |
| Preoperative Hb below 10 g/dL, n (%) | 41 (10.6) | 5 | 100 (13.3) | 47 |
| Past blood transfusion, n (%) | 50 (13.7) | 27 | 136 (18.0) | 45 |
| Abbreviations: Hb, hemoglobin; IQR, interquartile range. |  |  |  |  |
| Observed data before imputation; percentages use women with the item recorded as the denominator. |  |  |  |  |

In the development cohort, the OR for hysterectomy per additional score point was 1.90 (95% confidence interval [CI]: 1.60, 2.25), giving the development equation

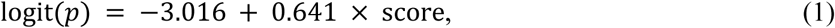

with a C-statistic of 0.76 (95% CI: 0.70, 0.80). There was no evidence of nonlinearity in the score (*p* = 0.247). In the eight-item model, older age (adjusted OR [aOR] = 1.20 per year; 95% CI: 1.13, 1.27), a uterus palpable at or above the umbilicus (aOR = 9.62; 95% CI: 1.94, 47.79), previous myomectomy (aOR = 5.84; 95% CI: 1.65, 20.68) and previous laparotomy (aOR = 3.25; 95% CI: 1.20, 8.82) showed the strongest associations with hysterectomy (**Table S2**).

### Validation and updating

In the validation cohort, the development equation retained its discrimination, with a C-statistic of 0.73 (95% CI: 0.69, 0.77), but overestimated risk; the O/E ratio was 0.69 (95% CI: 0.60, 0.78), the calibration intercept was −0.60 (95% CI: −0.79, −0.42) and the calibration slope was 0.83 (95% CI: 0.63, 1.03) (**Table 3**). The observed hysterectomy proportion was lower than the predicted risk in every score group, from 2.7% observed against 8.4% predicted at scores of 0 to 1 to 46.4% against 84.0% at scores of ≥7 (**Figures 3 and 4**).

**Figure 3.**
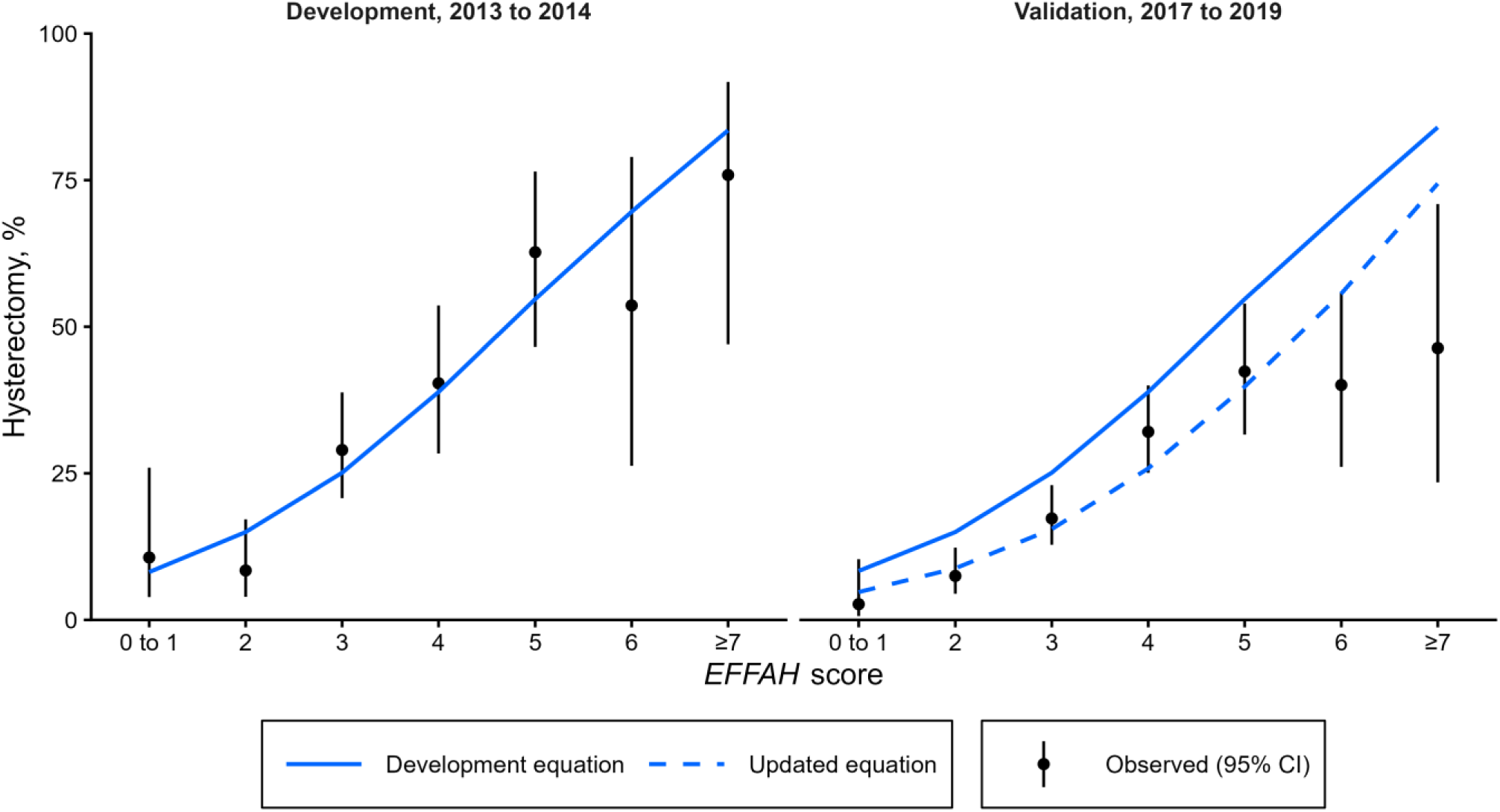
Observed hysterectomy (95% CI) and predicted risk by *EFFAH* score in the development and validation cohorts. Abbreviations: CI, confidence interval; *EFFAH*, Estimating Fibroid Factors Associated with Hysterectomy.

**Figure 4.**
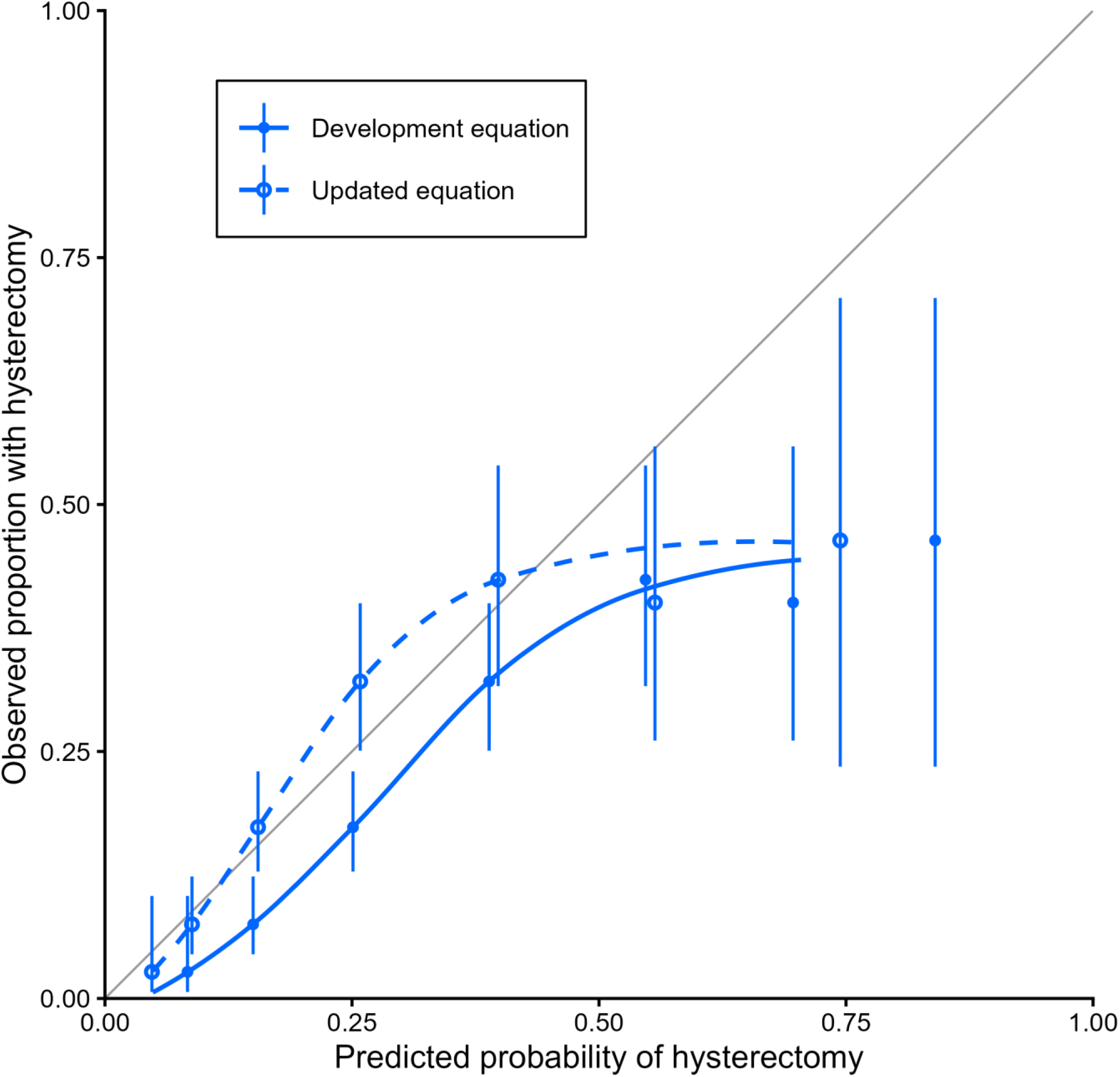
Calibration of the development and updated equations in the validation cohort, with observed proportions by score group (95% CI). Abbreviations: CI, confidence interval.

**Table 3.** Discrimination and calibration of the *EFFAH* score equation in development and validation.

| Measure | Development, apparent | Validation, development equation | Validation, updated equation (apparent) | Validation, updated equation (optimism-corrected) |
| --- | --- | --- | --- | --- |
| C-statistic (95% CI) | 0.76 (0.70, 0.80) | 0.73 (0.69, 0.77) | 0.73 (0.69, 0.77) | 0.72 |
| Calibration intercept (95% CI) | 0.00 (-0.24, 0.24) | -0.60 (-0.79, -0.42) | 0.00 (-0.18, 0.18) | 0.03 |
| Calibration slope (95% CI) | 1.00 (0.73, 1.27) | 0.83 (0.63, 1.03) | 0.83 (0.63, 1.03) | 0.78 |
| Observed-to-expected ratio (95% CI) | 1.00 (0.86, 1.16) | 0.69 (0.60, 0.78) | 1.00 (0.87, 1.14) | 1.02 |
| Brier score | 0.178 | 0.162 | 0.152 | 0.160 |
| Abbreviations: CI, confidence interval; *EFFAH*, Estimating Fibroid Factors Associated with Hysterectomy.<br>Development equation: $\text{logit}(p) = -3.016 + 0.641 \times \text{score}$. Updating model selected by closed testing: intercept update. Updated equation: $\text{logit}(p) = -3.619 + 0.641 \times \text{score}$. | | | | |

The joint test of the calibration intercept and slope rejected the development equation (*p* < 0.001), whereas the test of the slope alone did not (*p* = 0.100), so only the intercept was re-estimated, giving the updated equation

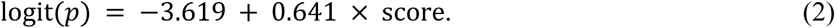

The intercept-only update was selected in 87% of bootstrap samples and the intercept and slope update in 13%. After correcting for optimism, the calibration slope of the updated equation was 0.78 and the O/E ratio 1.02. The updated equation still overestimated risk at the highest scores, where the adjusted risk at ≥7 was 74.4% against an observed proportion of 46.4% (**Figure 3**).

### Risk groups and clinical utility

The proportion with hysterectomy increased across the three risk groups in both cohorts (**Table 1**, **Figure 3**). In the development cohort, they were 9.1% (95% CI: 5.0, 15.9) in the low-risk group, 33.4% (95% CI: 26.4, 41.2) in the intermediate-risk group and 63.8% (95% CI: 52.3, 73.9) in the high-risk group. In the validation cohort, they were 6.2% (95% CI: 3.8, 9.9), 23.5% (95% CI: 19.5, 28.1) and 42.2% (95% CI: 34.2, 50.6), and the corresponding adjusted risks from the updated equation were 7.7%, 19.8% and 49.3% (**Figure 2**). About one third of validation records fell in the low-risk group. A score of ≥3 identified 90.3% (95% CI: 84.7, 94.0) of validation hysterectomies, and a score below 3 yielded a negative predictive value of 93.8% (95% CI: 90.1, 96.2). A score of ≥5 yielded a specificity of 86.7% (95% CI: 83.8, 89.2) and a positive predictive value of 42.2% (95% CI: 34.2, 50.6) (**Table S5**).

Using the development equation to identify women at high risk of hysterectomy gave a higher net benefit than treating all or no women as high risk at all threshold probabilities from 10% to 36% (**Figure 5**); at a threshold of 20%, the net benefit was 0.072, compared with 0.014 for treating all women as high risk. In the validation cohort, median estimated blood loss was 300 mL in the low-risk group, 389 mL in the intermediate-risk group and 329 mL in the high-risk group, and intraoperative transfusion was recorded for 1.8%, 3.7% and 5.6% of women in these groups; relaparotomy was infrequent in all groups (**Table S6**).

**Figure 5.**
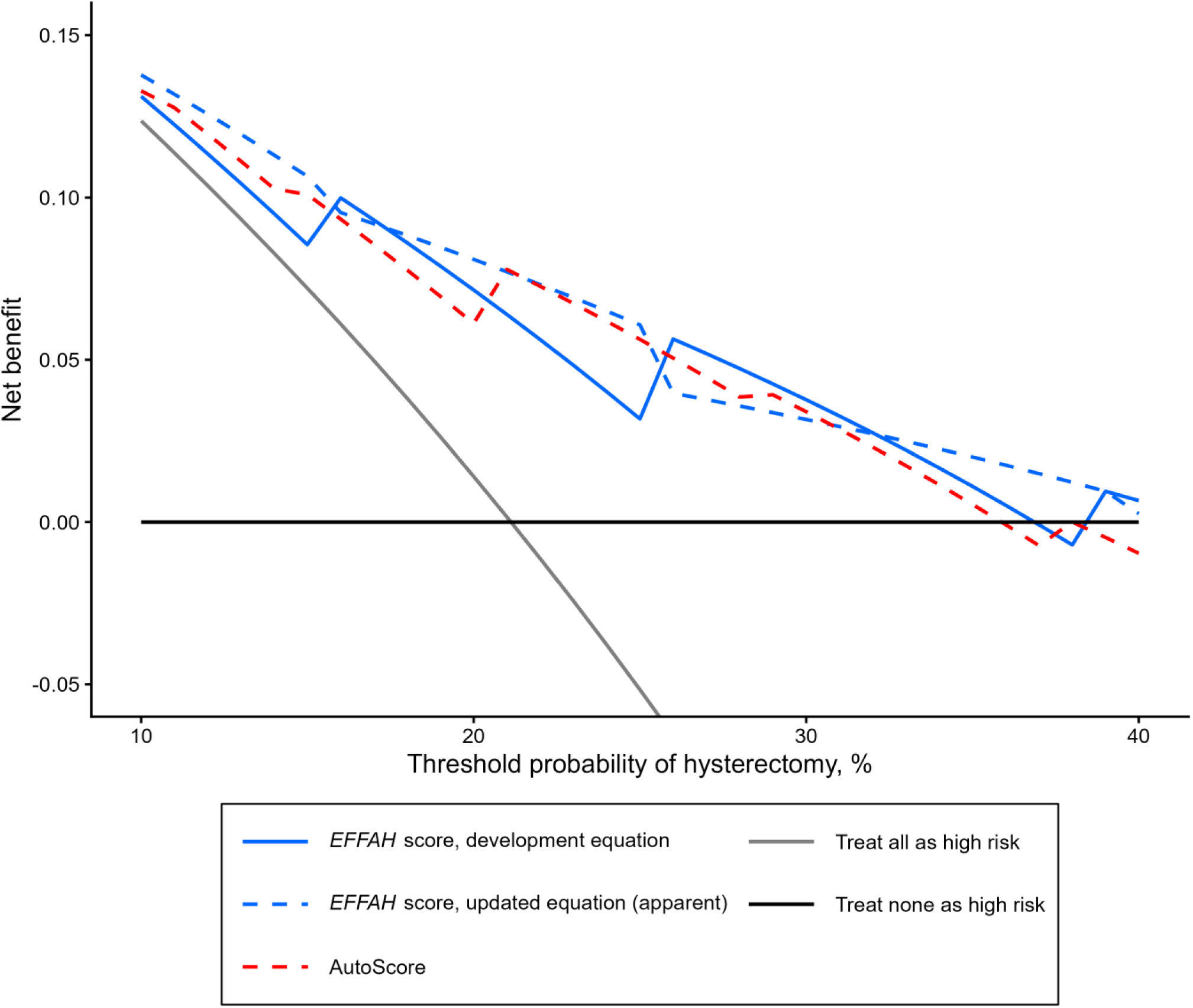
Net benefit of using the score (development and updated equations) and AutoScore to identify women at high risk of hysterectomy, by threshold probability, validation cohort. Abbreviations: *EFFAH*, Estimating Fibroid Factors Associated with Hysterectomy.

### AutoScore comparison and sensitivity analyses

AutoScore yielded a C-statistic of 0.74 (95% CI: 0.69, 0.78) in the validation cohort, which did not differ significantly from that of the *EFFAH* score (difference 0.01; 95% CI: −0.02, 0.04; *p* = 0.542). Its calibration was poorer, with a calibration slope of 0.62 (95% CI: 0.47, 0.78) and an O/E ratio of 0.66 (95% CI: 0.58, 0.75) (**Table S7**, **Figure 6**).

**Figure 6.**
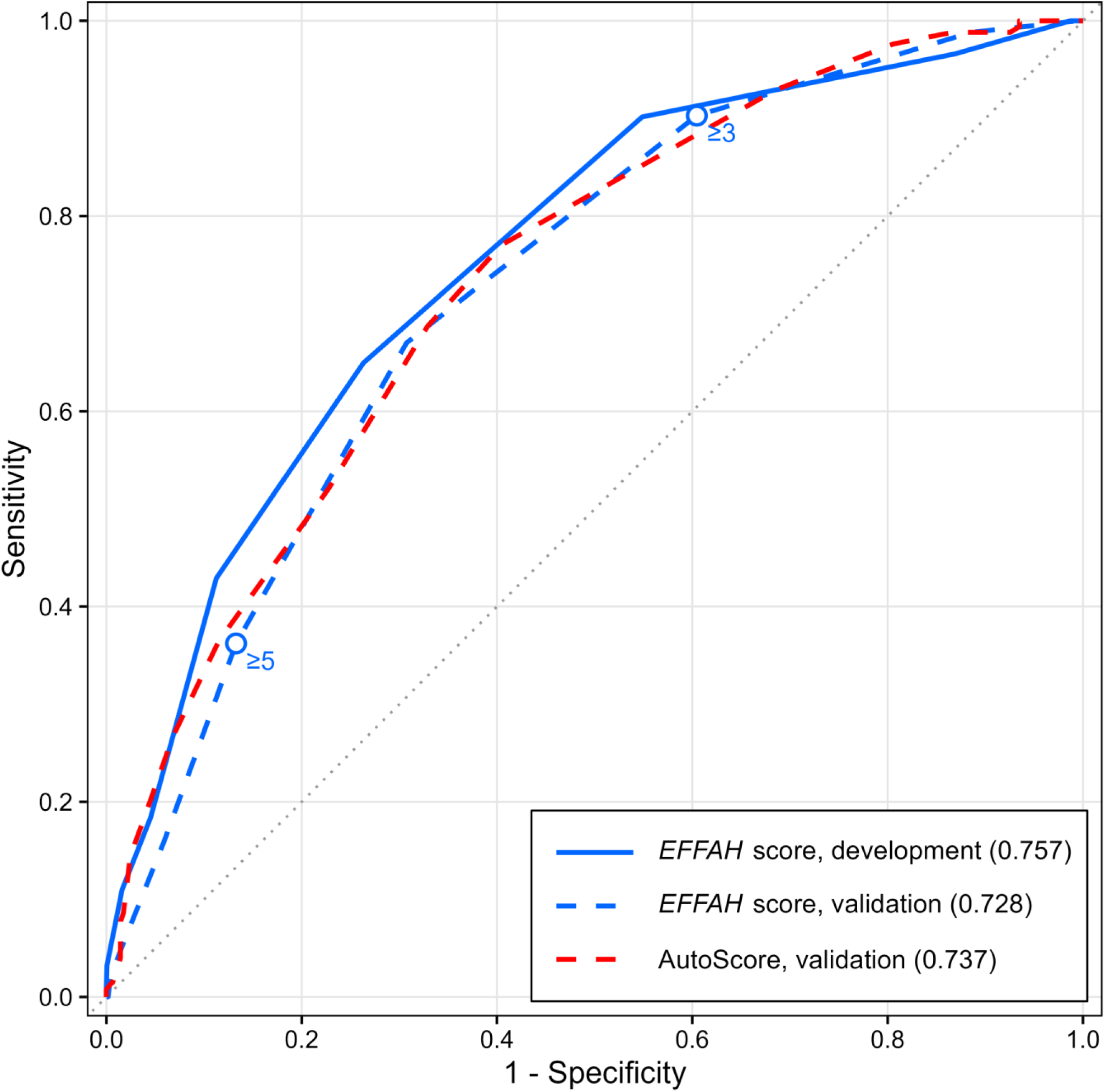
Receiver operating characteristic curves of the *EFFAH* score in both cohorts and of AutoScore in the validation cohort. C-statistics are shown in parentheses; circles mark the risk-group cut-offs at 3 and 5 points in the validation cohort. Abbreviations: *EFFAH*, Estimating Fibroid Factors Associated with Hysterectomy.

Scoring hemoglobin at first encounter yielded a C-statistic of 0.73 (95% CI: 0.69, 0.77) and a lower O/E ratio of 0.60 (95% CI: 0.53, 0.69). The complete-case analysis of 602 records gave a C-statistic of 0.72 (95% CI: 0.67, 0.76) and an O/E ratio of 0.67 (95% CI: 0.57, 0.78) (**Table S4**). With 800 records and 169 hysterectomies, the approximate 95% CI widths were 0.08 for the C-statistic, 0.27 for the O/E ratio and 0.39 for the calibration slope.

## Discussion

In a later cohort at the same district hospital, the *EFFAH* score ranked women counseled for myomectomy by their risk that the operation would end as hysterectomy almost as well as in the cohort in which its equation was fitted, and its three risk groups separated women at low, intermediate and high risk in both periods. What did not transfer in the validation was the absolute level of risk; fewer operations ended in hysterectomy in 2017 to 2019, so the development equation overstated the risk of women at every score, and re-estimating the intercept corrected this on average. A data-driven points score built from the same items with the AutoScore random forest approach discriminated no better and was less well calibrated.

The decreased proportion of women with hysterectomy between the two periods occurred although the validation cohort had, if anything, a slightly less favorable case mix. The shift therefore likely reflects a change in practice more than a change in the women operated on, for example in surgical experience with large fibroid uteri, operative technique or the threshold at which surgeons proceed to hysterectomy, none of which the score captures. Preserved discrimination with shifted calibration in the large is the expected pattern when a model is applied where the outcome frequency differs, and an intercept update is the recommended first correction^36,53^. In empirical comparisons, it performed as well as more extensive revision with validation samples of this size^54,55^.

Hysterectomy was far more frequent in both cohorts than in the United States series cited in the Introduction^18,19^. Two features of this study may explain the difference. The outcome in this study was hysterectomy among women counseled for myomectomy with possible hysterectomy, which includes hysterectomies decided during surgery based on operative findings and prior consent, not only hysterectomies necessitated by hemorrhage. The women also presented with large uteri, as in other sub-Saharan series^4,5^, and high hysterectomy frequencies have been reported elsewhere with unfavorable case mix: 6% of repeat abdominal myomectomies for large uteri in the UK^20^ and 8.4% of laparoscopic myomectomies in a Korean series^56^. The near-absence of conversion in the Nigerian series cited earlier may reflect their narrower outcome, unplanned conversion during myomectomy, not necessarily a lower underlying risk alone^7,14,22^.

Published tools most similar in format include points scores for transfusion at myomectomy from a United States registry: one with an area under the receiver operating characteristic curve (AUC) of 0.792 and bootstrap internal validation only^24^, and one with a preoperative AUC of 0.69 that its authors judged too low for clinical use^25^. An integer score for conversion of laparoscopic to open myomectomy from a single French center had a C-statistic of 0.87 without validation^26^. Externally validated models for conversion of laparoscopic to open hysterectomy in the UK had AUCs of 0.65 to 0.68 with calibration slopes of 0.97 to 0.98^27^. Recent machine learning classifiers of surgery type in fibroid disease reported hormone-based accuracy of 1.00 in a cohort not restricted to women seeking uterine preservation, without calibration or independent validation^57^. Another bedside score developed in 168 hospitals across Africa for severe postoperative complications, the African Surgical Outcomes Study surgical risk calculator^58^, had an AUC of 0.73 at external validation in Nigeria^59^. The discrimination of the *EFFAH* score lies within the range of these validated tools, and this study adds the calibration, updating and net benefit assessment that TRIPOD+AI requires^35^. Within this study, AutoScore offered no discriminative advantage, consistent with a systematic review that found no AUC benefit of machine learning over logistic regression in comparisons at low risk of bias^38^, and its poorer calibration in the new period probably reflects point weights estimated from a small development sample.

For counseling, the score turns routinely collected findings into a frequency that a woman can weigh before she consents. Guidance from Canada recommends counseling about the risk of hysterectomy at a planned myomectomy without providing an estimate^16^, guidance from the United States does not address it^17^, and consent guidance recommends presenting surgical risks as frequencies with verbal descriptors^60^. The adjusted risks of the three groups provide such frequencies. A low score supports reassurance, and a high score supports preparation for hysterectomy; because overestimation persisted at the highest scores, where few women contributed, risks at scores of ≥6 are better communicated as high than as precise percentages. Because calculation requires only addition, the items contributing to a woman’s risk are visible to her and to the surgeon.

The same score could serve different needs in different settings. Where resources are limited, women often present with large uteri^4^, myomectomy is usually performed by open surgery, and transfusion is more frequent^5–7^. In such cases, the score could direct resources before surgery, i.e., readily available cross-matched blood, a senior surgeon or referral could be reserved for women at high risk, while women at low risk, about one third of this cohort, could be operated on with standard preparation. Because it needs no magnetic resonance imaging, hormone assay or additional laboratory test, it can be applied wherever a history, an abdominal examination, a hemoglobin measurement and an abdominopelvic ultrasound are available. In high-resource settings, hysterectomy at a planned myomectomy is less frequent but has been described as an underestimated risk^18^. In these settings, the score could give consent a number and inform planning, such as the choice of surgeon or route, although imaging may add information that the score does not capture, and the lower baseline frequency means the intercept would need re-estimation.

At Catholic Hospital, Battor, the updated equation can inform counseling now. Elsewhere, in district hospitals and tertiary centers alike, the intercept should be re-estimated wherever the proportion with hysterectomy differs, and discrimination should be confirmed, because surgeons, case mix and the threshold for hysterectomy vary^61,62^. The next step is geographic validation in Ghana, the region and high-resource settings, ideally prospective, with the score recorded at the preoperative visit and the intended operation documented. Whether triage guided by the score changes transfusion, referral or the frequency of hysterectomy needs evaluation before it is adopted clinically. Future studies may examine outcomes beyond perioperative complications in women with high *EFFAH* scores whose operations end as myomectomy instead of hysterectomy.

### Strengths and limitations

In terms of strengths, the *EFFAH* score points were taken from the scoring form and were not re-estimated, and the validation sample size was fixed from the pilot before the validation records were extracted, so the achieved precision can be judged against the criteria proposed for external validation^44,45^. Calibration was also assessed across the calibration hierarchy^48^ and the updating method was selected by closed testing (instead of by inspection)^49^.

The study, however, has several limitations. First, this is a single-center temporal validation, and discrimination and calibration in other hospitals, where surgeons, case mix and the threshold for hysterectomy differ, are unknown^61,62^. Surgeons, operative protocols and blood availability were not recorded for the development cohort, so changes in the setting between the two periods could not be described. Why a woman wished to keep her uterus (whether for future childbearing or for religious or other personal reasons) was not recorded; age and parity reflect it only indirectly, and it may change how readily the surgeon proceeds to hysterectomy. Second, the risk-group cut-offs were informed by the observed proportions, so the proportions within each group are optimistic and the cut-offs at 3 and 5 points require confirmation in new data. Third, the updated equation was evaluated in the cohort used to update it, so its performance estimates are apparent, although bootstrap optimism was small. Fourth, renal ultrasound was missing for almost half of development records and was multiply imputed, even though the complete-case analysis gave similar discrimination. Fifth, AutoScore was trained on about 235 records (60% of one imputed development dataset), and an advantage for machine learning models trained on larger datasets or additional variables cannot be excluded.

## Conclusion

Among women with leiomyomas counseled for myomectomy with possible hysterectomy at a district hospital in Ghana, the *EFFAH* score (an eight-item bedside score built from routine preoperative findings) ranked women by their risk of hysterectomy in a later cohort, and an intercept update aligned its predicted risks with the observed proportion. A data-driven alternative built with AutoScore offered no improvement. The score gives women and surgeons an individual risk estimate to discuss at consent and could guide the use of blood, senior surgeons and referral where these are scarce. Because it requires no imaging beyond ultrasound and no laboratory test beyond hemoglobin, it could become a common basis for counseling in resource-limited and resource-rich settings alike, once its intercept has been re-estimated locally and its performance confirmed in external validation.

## Declarations

### Ethics approval

Ethical clearance was granted by the Ethics Committee of Catholic Hospital, Battor (approval number CHB-ERC-002/01/24). The study was performed in accordance with the 1964 Declaration of Helsinki and its later amendments. All information was anonymized and stored on a password-protected computer.

### Consent to participate

The requirement for individual informed consent was waived by the Ethics Committee of Catholic Hospital, Battor, because the study used anonymized records collected during routine care.

### Consent for publication

Not applicable.

## Data Availability

The data and analysis code underlying this work are available from the corresponding author on reasonable request with the permission of Catholic Hospital, Battor.

## Acknowledgements

The authors thank all the teams and health workers involved in the care of the women in this study, and the Information Technology Department of the Cervical Cancer Prevention and Training Centre, Catholic Hospital, Battor, for assistance with data compilation.

## CRediT authorship contribution statement

Conceptualization: KE and NOME. Methodology: KE and NOME. Investigation (data collection): BHA, AKA, CN, SD, EYL, HVD, AAA, FA, JOP, EOKA, NOME and KE. Data curation: NOME, JEA, AKA, SD and KE. Formal analysis: NOME, JEA, AKA, SD and KE. Writing, original draft: NOME, JEA and KE. All authors read and approved the final manuscript.

## References

1. Wu, C. et al. The epidemiology of uterine fibroids: global disease burden from 1990 to 2021 and future trend predictions. Frontiers in reproductive health 7, 1629834 (2025).

2. Munro, M. G., Tchaikovski, S. N. & Murji, A. The epidemiology and pathogenesis of uterine fibroids. Int. J. Gynaecol. Obstet. 171, 1029–1045 (2025).

3. Stewart, E. A., Cookson, C. L., Gandolfo, R. A. & Schulze-Rath, R. Epidemiology of uterine fibroids: a systematic review. BJOG 124, 1501–1512 (2017).

4. Adegbesan-Omilabu, M. A., Okunade, K. S. & Gbadegesin, A. Knowledge of, Perception of, and Attitude towards Uterine Fibroids among Women with Fibroids in Lagos, Nigeria. Scientifica (Cairo). 2014, 809536 (2014).

5. Edema, R. O., Okonta, P., Adeyinka, A., Ekoh, A. & Jagu, V. L. Effectiveness of tourniquet with and without adjunctive misoprostol on haemorrhage during open myomectomy: A randomised controlled trial. Afr. J. Reprod. Health. (Afr J Reprod Health 2026; 30 [13]: 33–44). (2026) doi:10.29063/ajrh2026/v30i13.4.

6. Ogwu, R. O. et al. Tourniquet with Intravenous Tranexamic Acid versus Tourniquet with Vaginal Misoprostol for Blood Loss Reduction during Open Myomectomy: A Double-Blind Randomised Trial. Niger. Med. J. 67, 1530–1540 (2026).

7. Adesina, K. T., Owolabi, B. O., Raji, H. O. & Olarinoye, A. O. Abdominal myomectomy: A retrospective review of determinants and outcomes of complications at the University of Ilorin Teaching Hospital, Ilorin, Nigeria. Malawi Med. J. 29, 37–42 (2017).

8. Igboeli, P., Walker, W., McHugh, A., Sultan, A. & Al-Hendy, A. Burden of Uterine Fibroids: An African Perspective, A Call for Action and Opportunity for Intervention. Curr. Opin. Gynecol. Obstet. 2, 287–294 (2019).

9. Okeke, T. C., Okezie, O. A., Obioha, K. C. E., Ikeako, L. C. & Ezenyeaku, C. C. Trends of myomectomy at the University of Nigeria Teaching Hospital (UNTH) Enugu Nigeria. Niger. J. Med. 20, 224–7 (2011).

10. Omole-Ohonsi, A. & Ashimi, O. A. Non-emergency hysterectomy: why the aversion? Arch. Gynecol. Obstet. 280, 953–9 (2009).

11. Alkhrait, S., Malasevskaia, I. & Madueke-Laveaux, O. S. Fibroids and Fertility. Obstet. Gynecol. Clin. North Am. 50, 663–675 (2023).

12. Frisch, E. H. et al. The Impact of Fertility Goals on Long-term Quality of Life in Reproductive-aged Women Who Underwent Myomectomy versus Hysterectomy for Uterine Fibroids. J. Minim. Invasive Gynecol. 30, 642–651 (2023).

13. Wolgemuth, T., Suresh, N., Holder, K. & Milad, M. P. Informational Preferences of Women Seeking Treatment for Symptomatic Uterine Fibroids: A Qualitative Study. J. Minim. Invasive Gynecol. 30, 284–289 (2023).

14. Omole-Ohonsi, A. & Belga, F. Surgical management of uterine fibroids at aminu kano teaching hospital. Obstet. Gynecol. Int. 2012, 702325 (2012).

15. Igbodike, E. et al. Giant ‘Hydra Headed’ Uterine Fibroid in a Nullipara: A Case Report. Clin. Med. Insights Case Rep. 17, 11795476241274688 (2024).

16. Vilos, G. A., Allaire, C., Laberge, P.-Y., Leyland, N. & SPECIAL CONTRIBUTORS. The management of uterine leiomyomas. J. Obstet. Gynaecol. Can. 37, 157–178 (2015).

17. Management of Symptomatic Uterine Leiomyomas: ACOG Practice Bulletin, Number 228. Obstetrics and gynecology 137, e100–e115 (2021).

18. Coyne, K., Purdy, M. P., Bews, K. A., Habermann, E. B. & Khan, Z. Risk of hysterectomy at the time of myomectomy: an underestimated surgical risk. Fertil. Steril. 121, 107–116 (2024).

19. Dallas, K. et al. Racial Disparities in Outcomes of Women Undergoing Myomectomy. Obstetrics and gynecology 138, 845–851 (2021).

20. Teo, U. L., Kopeika, J., Pundir, J. & El-Toukhy, T. Peri-operative morbidity and fertility outcome after repeat abdominal myomectomy for large fibroid uterus. J. Obstet. Gynaecol. 40, 673–677 (2020).

21. Coyne, K., Purdy, M. P., Bews, K. A., Habermann, E. B. & Khan, Z. Reply of the authors: comment on ‘Risk of hysterectomy at the time of myomectomy - An underestimated surgical risk’. Fertil. Steril. 121, 134 (2024).

22. Agboola, A. D., Bello, O. O. & Olayemi, O. O. A clinical audit of the patterns of presentations and complications of abdominal myomectomy at the University College Hospital, Ibadan, Nigeria. J. Obstet. Gynaecol. 41, 1145–1150 (2021).

23. Oladapo, O. T. & Akinsanya, A. F. Relative morbidity of abdominal myomectomy for very large uterine fibroids in a developing country hospital. Arch. Gynecol. Obstet. 283, 825–30 (2011).

24. Klebanoff, J. S. et al. A Clinically Applicable Prediction Model for the Risk of Transfusion in Women Undergoing Myomectomy. J. Minim. Invasive Gynecol. 28, 1765–1773.e1 (2021).

25. Hamilton, K. M. et al. Characteristics associated with blood transfusion among women undergoing laparoscopic myomectomy: a National Surgical Quality Improvement Program study. Am. J. Obstet. Gynecol. 231, 109.e1–109.e9 (2024).

26. Dubuisson, J. B. et al. Laparoscopic myomectomy: predicting the risk of conversion to an open procedure. Hum. Reprod. 16, 1726–31 (2001).

27. Madhvani, K., Fernandez-Felix, B. M., Zamora, J., Carpenter, T. & Khan, K. S. Personalising the risk of conversion from laparoscopic to open hysterectomy in benign conditions: Development and external validation of risk prediction models. BJOG 129, 1141–1150 (2022).

28. Madhvani, K. et al. Predicting major complications in patients undergoing laparoscopic and open hysterectomy for benign indications. CMAJ 194, E1306–E1317 (2022).

29. DeLong, A. et al. Individualized Assessment of Risk of Complications after Benign Hysterectomy. J. Minim. Invasive Gynecol. 29, 976–983 (2022).

30. Roth, T. M., Gustilo-Ashby, T., Barber, M. D. & Myers, E. R. Effects of race and clinical factors on short-term outcomes of abdominal myomectomy. Obstetrics and gynecology 101, 881–4 (2003).

31. Murji, A. et al. Risks of preoperative anemia in women undergoing elective hysterectomy and myomectomy. Am. J. Obstet. Gynecol. 221, 629.e1–629.e18 (2019).

32. Antunes, D., Gante, I., Carvalho, M. J., Medeiros-Borges, C. & Águas, F. The impact of anemia on treatment management and clinical outcomes of women hospitalized for uterine leiomyomas. Ginekol. Pol. 93, 799–805 (2022).

33. Pundir, J. et al. Peri-operative morbidity associated with abdominal myomectomy for very large fibroid uteri. Eur. J. Obstet. Gynecol. Reprod. Biol. 167, 219–24 (2013).

34. Amisu, M., Chionuma, J., Odeyemi, A. & Odetayo, O. Prevalence and pattern of renal complications of uterine fibroids in a Teaching Hospital in Nigeria. Afr. Health Sci. 24, 156–162 (2024).

35. Collins, G. S. et al. TRIPOD+AI statement: updated guidance for reporting clinical prediction models that use regression or machine learning methods. BMJ 385, e078378 (2024).

36. Van Calster, B. et al. Calibration: the Achilles heel of predictive analytics. BMC Med. 17, 230 (2019).

37. Vickers, A. J., Van Calster, B. & Steyerberg, E. W. Net benefit approaches to the evaluation of prediction models, molecular markers, and diagnostic tests. BMJ 352, i6 (2016).

38. Christodoulou, E. et al. A systematic review shows no performance benefit of machine learning over logistic regression for clinical prediction models. J. Clin. Epidemiol. 110, 12–22 (2019).

39. Dunyo, P., Effah, K. & Udofia, E. A. Factors associated with late presentation of cervical cancer cases at a district hospital: a retrospective study. BMC Public Health 18, 1156 (2018).

40. Effah, K. et al. Expanding management strategies for cervical precancerous lesions in resource-limited settings: insights from a training center in a district hospital in Ghana. BMC Womens Health 24, 428 (2024).

41. Effah, K. et al. Nationwide cervical precancer screening in Ghana: concurrent HPV DNA testing and visual inspection under an expanded hub-and-spoke model. Sci. Rep. 15, 1744 (2025).

42. Krings, A. et al. Characterization of Human Papillomavirus prevalence and risk factors to guide cervical cancer screening in the North Tongu District, Ghana. PLoS One 14, e0218762 (2019).

43. Faul, F., Erdfelder, E., Buchner, A. & Lang, A.-G. Statistical power analyses using G*Power 3.1: tests for correlation and regression analyses. Behav. Res. Methods 41, 1149–60 (2009).

44. Riley, R. D. et al. Minimum sample size for external validation of a clinical prediction model with a binary outcome. Stat. Med. 40, 4230–4251 (2021).

45. Riley, R. D. et al. Evaluation of clinical prediction models (part 3): calculating the sample size required for an external validation study. BMJ 384, e074821 (2024).

46. White, I. R., Royston, P. & Wood, A. M. Multiple imputation using chained equations: Issues and guidance for practice. Stat. Med. 30, 377–99 (2011).

47. Sterne, J. A. C. et al. Multiple imputation for missing data in epidemiological and clinical research: potential and pitfalls. BMJ 338, b2393 (2009).

48. Van Calster, B. et al. A calibration hierarchy for risk models was defined: from utopia to empirical data. J. Clin. Epidemiol. 74, 167–76 (2016).

49. Vergouwe, Y. et al. A closed testing procedure to select an appropriate method for updating prediction models. Stat. Med. 36, 4529–4539 (2017).

50. Vickers, A. J. & Elkin, E. B. Decision curve analysis: a novel method for evaluating prediction models. Med. Decis. Making 26, 565–74 (2006).

51. Van Calster, B. et al. Reporting and Interpreting Decision Curve Analysis: A Guide for Investigators. Eur. Urol. 74, 796–804 (2018).

52. Xie, F., Chakraborty, B., Ong, M. E. H., Goldstein, B. A. & Liu, N. AutoScore: A Machine Learning-Based Automatic Clinical Score Generator and Its Application to Mortality Prediction Using Electronic Health Records. JMIR Med. Inform. 8, e21798 (2020).

53. Ramspek, C. L., Jager, K. J., Dekker, F. W., Zoccali, C. & van Diepen, M. External validation of prognostic models: what, why, how, when and where? Clin. Kidney J. 14, 49–58 (2020).

54. Janssen, K. J. M., Moons, K. G. M., Kalkman, C. J., Grobbee, D. E. & Vergouwe, Y. Updating methods improved the performance of a clinical prediction model in new patients. J. Clin. Epidemiol. 61, 76–86 (2008).

55. Steyerberg, E. W., Borsboom, G. J. J. M., van Houwelingen, H. C., Eijkemans, M. J. C. & Habbema, J. D. F. Validation and updating of predictive logistic regression models: a study on sample size and shrinkage. Stat. Med. 23, 2567–86 (2004).

56. Yoo, E. H. & Lee, S. K. Factors affecting completion of laparoscopic myomectomy. Clin. Exp. Obstet. Gynecol. 40, 78–80 (2013).

57. Öz, İ. & Öz, A. U. Prediction of surgery type for uterine fibroids using machine learning algorithms and hormone values. Medicine 105, e47965 (2026).

58. Kluyts, H.-L. et al. The ASOS Surgical Risk Calculator: development and validation of a tool for identifying African surgical patients at risk of severe postoperative complications. Br. J. Anaesth. 121, 1357–1363 (2018).

59. Osinaike, B. et al. Nigerian surgical outcomes - Report of a 7-day prospective cohort study and external validation of the African surgical outcomes study surgical risk calculator. Int. J. Surg. 68, 148–156 (2019).

60. Topcu, E. G. et al. FIGO best practice guidance in surgical consent. Int. J. Gynaecol. Obstet. 163, 795–812 (2023).

61. Collins, G. S. et al. Evaluation of clinical prediction models (part 1): from development to external validation. BMJ 384, e074819 (2024).

62. Debray, T. P. A. et al. A new framework to enhance the interpretation of external validation studies of clinical prediction models. J. Clin. Epidemiol. 68, 279–89 (2015).

