## Supplementary Information (Tables S1 to S7, Figures S1 and S2) for "Anticipating conversion to hysterectomy in women counseled for myomectomy: temporal validation of the *EFFAH* score at a Ghanaian district hospital"

**Table S1. Missing data and imputation diagnostics in the development (n = 392) and validation (n = 800) cohorts.**

| Item | Development missing, n (%) | Validation missing, n (%) | Validation imputation method |
|---|---|---|---|
| Age | 0 (0.0) | 0 (0.0) | none (complete) |
| Parity | 10 (2.6) | 35 (4.4) | pmm |
| Abdominal examination | 18 (4.6) | 19 (2.4) | polyreg |
| Previous myomectomy | 22 (5.6) | 15 (1.9) | logreg |
| Previous laparotomy | 18 (4.6) | 11 (1.4) | logreg |
| Renal ultrasound | 177 (45.2) | 142 (17.8) | polyreg |
| Preoperative hemoglobin | 5 (1.3) | 47 (5.9) | pmm |
| Past transfusion | 27 (6.9) | 45 (5.6) | logreg |
| C-statistic, fraction of missing information | 0.07 | 0.03 | |
| C-statistic, Monte Carlo standard error | 0.0014 | 0.0007 | |
| Calibration intercept, fraction of missing information | 0.02 | 0.01 | |
| Calibration intercept, Monte Carlo standard error | 0.0033 | 0.0020 | |
| Calibration slope, fraction of missing information | 0.04 | 0.05 | |
| Calibration slope, Monte Carlo standard error | 0.0055 | 0.0048 | |

Abbreviations: logreg, logistic regression; pmm, predictive mean matching; polyreg, polytomous regression.

**Table S2. Logistic regression of hysterectomy in the development cohort, 20 imputations (n = 392).**

| Model | Predictor | OR (95% CI) | *p* value |
|---|---|---|---|
| Eight items | Age, per year | 1.20 (1.13, 1.27) | <0.001 |
| Eight items | Parity, per birth | 1.33 (1.00, 1.77) | 0.049 |
| Eight items | Uterus palpable below umbilicus (vs not palpable) | 5.93 (1.17, 30.16) | 0.032 |
| Eight items | Uterus palpable at or above umbilicus (vs not palpable) | 9.62 (1.94, 47.79) | 0.006 |
| Eight items | Previous myomectomy | 5.84 (1.65, 20.68) | 0.006 |
| Eight items | Previous laparotomy other than myomectomy | 3.25 (1.20, 8.82) | 0.021 |
| Eight items | Unilateral hydronephrosis (vs normal) | 3.82 (0.96, 15.24) | 0.058 |
| Eight items | Bilateral hydronephroses (vs normal) | 1.18 (0.21, 6.65) | 0.852 |
| Eight items | Preoperative Hb, per g/dL | 1.06 (0.87, 1.28) | 0.558 |
| Eight items | Past blood transfusion | 1.76 (0.86, 3.60) | 0.120 |
| Total score | *EFFAH* score, per point | 1.90 (1.60, 2.25) | <0.001 |

Abbreviations: aOR, adjusted odds ratio; CI, confidence interval; *EFFAH*, Estimating Fibroid Factors Associated with Hysterectomy; Hb, hemoglobin; OR, odds ratio.
Eight-item rows are adjusted odds ratios from a model including all eight items; the total-score row is an odds ratio from a model with the score alone.

**Table S3. Linearity check and closed-testing update of the *EFFAH* equation to the validation cohort.**

| Step | Result |
|---|---|
| Linearity of the score (development) | Restricted cubic spline, knots 1, 3, 5; likelihood ratio test $p$ = 0.247 for nonlinearity |
| Development equation | logit($p$) = -3.016 + 0.641 x score |
| Closed test 1: intercept and slope jointly (0, 1) | D = 22.14 on 2 and 40724 df; $p$ <0.001 |
| Closed test 2: slope equal to 1 | D = 2.70 on 1 and 5339 df; p = 0.100 |
| Selected update | intercept update |
| Updated equation | logit($p$) = -3.619 + 0.641 x score |
| Bootstrap selection frequency, %: no update / intercept / intercept and slope | 0.0 / 86.8 / 13.2 |
| Optimism in C-statistic | 0.012 |
| Optimism in calibration slope | 0.053 |
| Abbreviations: df, degrees of freedom; *EFFAH*, Estimating Fibroid Factors Associated with Hysterectomy. | |

**Table S4. Sensitivity analyses of *EFFAH* score performance with the development equation.**

| Cohort | Analysis | n (events) | C-statistic (95% CI) | Calibration intercept (95% CI) | Calibration slope (95% CI) | Observed-to-expected ratio (95% CI) |
|---|---|---|---|---|---|---|
| Development | Primary: 20 imputations | 392 (124) | 0.76 (0.70, 0.80) | 0.00 (-0.24, 0.24) | 1.00 (0.73, 1.27) | 1.00 (0.86, 1.16) |
| Development | Complete cases | 198 (54) | 0.71 (0.63, 0.78) | -0.22 (-0.56, 0.12) | 0.82 (0.46, 1.18) | 0.88 (0.70, 1.10) |
| Validation | Primary: preoperative Hb, 20 imputations | 800 (169) | 0.73 (0.69, 0.77) | -0.60 (-0.79, -0.42) | 0.83 (0.63, 1.03) | 0.69 (0.60, 0.78) |
| Validation | Hb at first encounter | 800 (169) | 0.73 (0.69, 0.77) | -0.84 (-1.02, -0.65) | 0.83 (0.64, 1.02) | 0.60 (0.53, 0.69) |
| Validation | Complete cases | 602 (121) | 0.72 (0.67, 0.76) | -0.64 (-0.85, -0.42) | 0.81 (0.59, 1.04) | 0.67 (0.57, 0.78) |

Abbreviations: CI, confidence interval; *EFFAH*, Estimating Fibroid Factors Associated with Hysterectomy; Hb, hemoglobin.
Complete cases: women with all eight items recorded.

**Table S5. Sensitivity, specificity and predictive values of *EFFAH* risk-group cut-offs.**

| Cut-off | Measure | Development, % (95% CI) | Development denominator, mean n | Development complete cases, k/n; % (95% CI) | Validation, % (95% CI) | Validation denominator, mean n | Validation complete cases, k/n; % (95% CI) |
|---|---|---|---|---|---|---|---|
| Score ≥3 (not low risk) | Sensitivity | 90.2 (82.9, 94.6) | 124 | 48/54; 88.9 (77.8, 94.8) | 90.3 (84.7, 94.0) | 169 | 108/121; 89.3 (82.5, 93.6) |
| Score ≥3 (not low risk) | Specificity | 45.1 (38.9, 51.5) | 268 | 55/144; 38.2 (30.7, 46.3) | 39.5 (35.7, 43.5) | 631 | 193/481; 40.1 (35.8, 44.6) |
| Score ≥3 (not low risk) | PPV | 43.2 (37.2, 49.4) | 259 | 48/137; 35.0 (27.6, 43.3) | 28.6 (24.9, 32.6) | 534 | 108/396; 27.3 (23.1, 31.9) |
| Score ≥3 (not low risk) | NPV | 90.9 (84.1, 95.0) | 133 | 55/61; 90.2 (80.2, 95.4) | 93.8 (90.1, 96.2) | 266 | 193/206; 93.7 (89.5, 96.3) |
| Score ≥5 (high risk) | Sensitivity | 42.9 (33.8, 52.5) | 124 | 21/54; 38.9 (27.0, 52.2) | 36.2 (29.1, 44.0) | 169 | 41/121; 33.9 (26.1, 42.7) |
| Score ≥5 (high risk) | Specificity | 88.7 (84.1, 92.1) | 268 | 128/144; 88.9 (82.7, 93.0) | 86.7 (83.8, 89.2) | 631 | 420/481; 87.3 (84.0, 90.0) |
| Score ≥5 (high risk) | PPV | 63.8 (52.3, 73.9) | 83 | 21/37; 56.8 (40.9, 71.3) | 42.2 (34.2, 50.6) | 145 | 41/102; 40.2 (31.2, 49.9) |
| Score ≥5 (high risk) | NPV | 77.1 (71.9, 81.5) | 309 | 128/161; 79.5 (72.6, 85.0) | 83.5 (80.5, 86.2) | 655 | 420/500; 84.0 (80.5, 87.0) |

Abbreviations: CI, confidence interval; *EFFAH*, Estimating Fibroid Factors Associated with Hysterectomy; NPV, negative predictive value; PPV, positive predictive value.
Complete cases: women with all eight items recorded. Complete-case 95% CIs use the Wilson score method.

**Table S6. Blood loss, intraoperative transfusion and relaparotomy by *EFFAH* risk group.**

| Cohort | Risk group | Women, n | Estimated blood loss, mL; median (IQR) | Blood loss recorded, n | Intraoperative transfusion, n/recorded | Relaparotomy, n/recorded |
|---|---|---|---|---|---|---|
| Development | Low (0 to 2) | 133 | 304 (217 to 414) | 51 | 1.4/65 | 0.0/127 |
| Development | Intermediate (3 to 4) | 175 | 366 (275 to 501) | 99 | 6.1/120 | 3.0/168 |
| Development | High (≥5) | 83 | 338 (250 to 401) | 63 | 5.5/65 | 1.0/74 |
| Validation | Low (0 to 2) | 266 | 300 (200 to 400) | 247 | 4.6/261 | 6.0/258 |
| Validation | Intermediate (3 to 4) | 389 | 389 (269 to 500) | 355 | 14.3/383 | 4.0/375 |
| Validation | High (≥5) | 145 | 329 (253 to 496) | 138 | 8.0/142 | 2.0/131 |

Abbreviations: *EFFAH*, Estimating Fibroid Factors Associated with Hysterectomy; IQR, interquartile range.
Counts are means across 20 imputations and can be fractional. Blood loss is the surgeon's estimate recorded in the theater register.

**Table S7. External performance of the *EFFAH* score and AutoScore, validation cohort (n = 800).**

| Model | C-statistic (95% CI) | Calibration intercept (95% CI) | Calibration slope (95% CI) | Observed-to-expected ratio (95% CI) | Difference in C-statistic vs *EFFAH* (95% CI); *p* |
|---|---|---|---|---|---|
| *EFFAH* score, development equation | 0.73 (0.69, 0.77) | -0.60 (-0.79, -0.42) | 0.83 (0.63, 1.03) | 0.69 (0.60, 0.78) | Reference |
| AutoScore points score | 0.74 (0.69, 0.78) | -0.74 (-0.94, -0.55) | 0.62 (0.47, 0.78) | 0.66 (0.58, 0.75) | 0.01 (-0.02, 0.04); *p* = 0.542 |

Abbreviations: CI, confidence interval; *EFFAH*, Estimating Fibroid Factors Associated with Hysterectomy.
Development performance of AutoScore is not reported because the development imputations used all development outcomes.

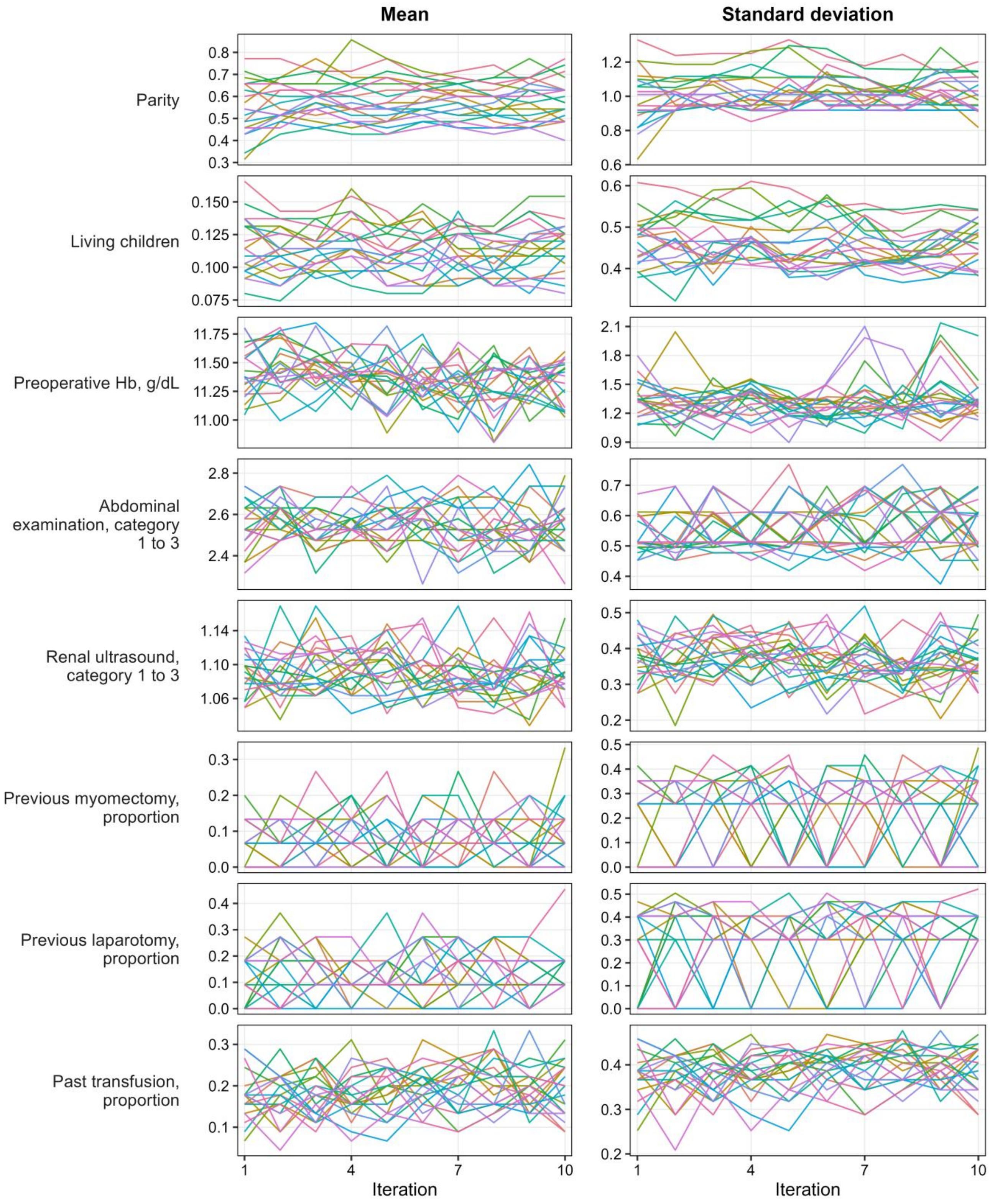


**Figure S1. Trace plots of the mean and standard deviation of imputed values by iteration for each imputed item in the validation cohort.**

Each line is one of 20 imputation chains. Previous laparotomy, missing for one record, is not shown.

Abbreviations: Hb, hemoglobin.

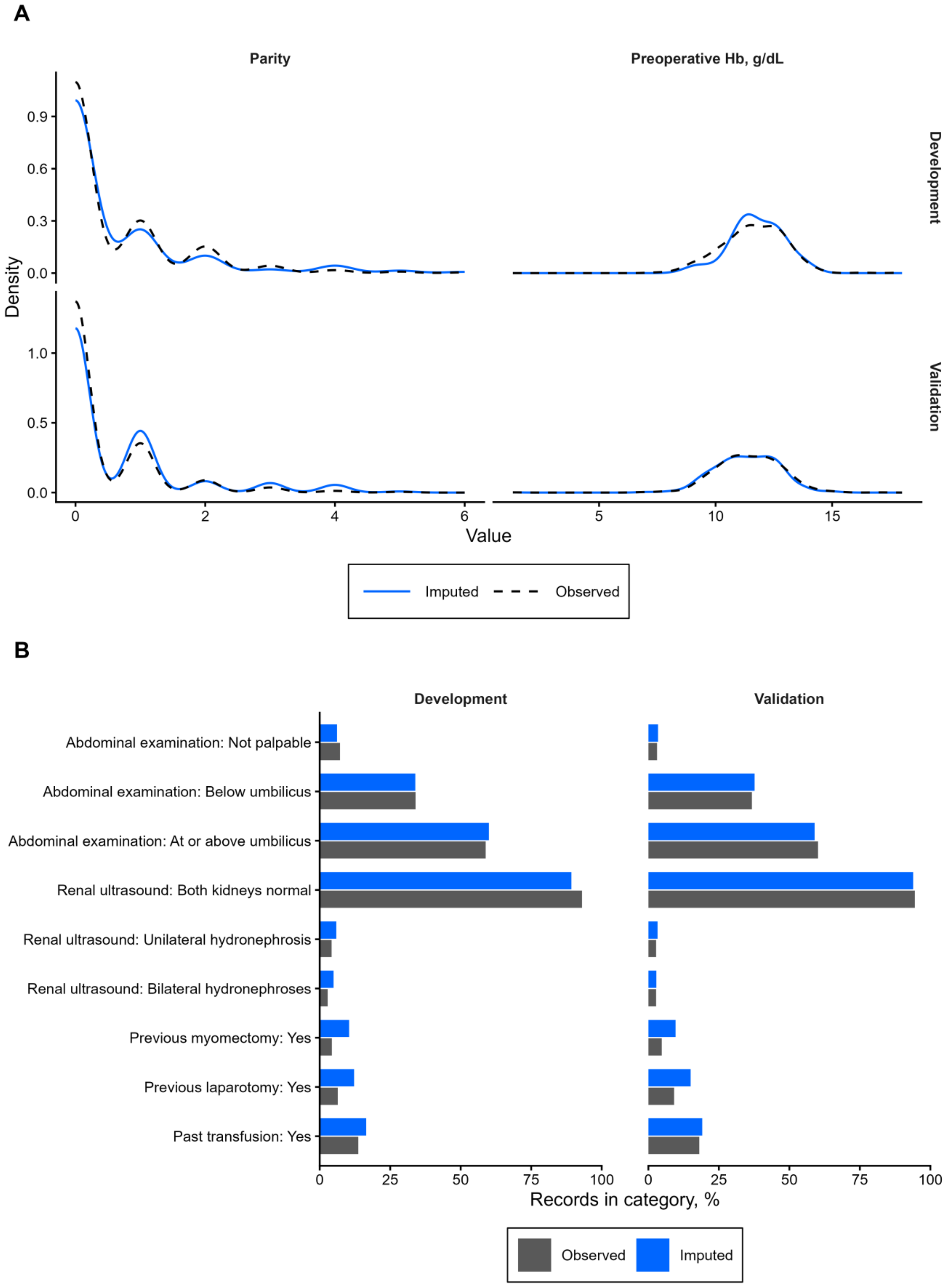


**Figure S2. Observed and imputed values in the development and validation cohorts. (A) Densities of parity and preoperative hemoglobin. (B) Proportions in each category of the categorical items.**

Abbreviations: Hb, hemoglobin.
